# Increase in Surgical Case Mix Index Across a Multi-Hospital System with Structured Limited Surgeon Chart Review

**DOI:** 10.64898/2026.10.13.26365430

**Authors:** Tovy Haber Kamine, Jacques Townsend, Samir M. Fakhry, Nicolas Jabbour, Edward Kelly

## Abstract

**Introduction:** Hospitals face increasing financial pressures. Hospital reimbursement is tied to the Case Mix Index (CMI). The CMI is a measure of complexity of care provided by the hospital and the total reimbursement is the CMI multiplied by a base pay factor, multiplied by the total number of discharges from the hospital. The few studies in the literature have involved logistically complex attempts to improve documentation. We sought to determine whether limited, structured chart review by surgeons can improve CMI in a multihospital system.

**Methods:** We performed a prospective, observational study in a four-hospital academic medical system on all surgical patients from 10/2022 to 7/2026. Two clinically busy trauma surgeons performed limited structured chart reviews for select surgical patients. In the limited intervention period (1/2024-9/2024) this focused only on the trauma division. In the full intervention period (10/2024-7/2026) the review was expanded to all surgical patients across the system. The primary outcome was change in surgical case mix index (CMI) over time.

**Results:** A total of 45,559 patients were included (13,807pre-intervention, 9,299 limited intervention, 22,453 full intervention). CMI increased significantly among surgical patients across the whole system (2.83 (95%CI 279-2.87) to 2.86 (95%CI 2.81-2.91) to 2.96 (95%CI 2.93-3.00), ANOVA p <0.001). Overall length of stay was unchanged among surgical patients across the whole system. The estimated increase in revenue per patient attributable to this intervention was $1,765 in surgical patients across the system.

**Conclusion:** Having trauma surgeons review targeted surgical patients for high value documentation capture significantly enhanced the CMI and estimated revenue of surgical services across a multi-hospital integrated health system. Such documentation reviews may represent a practical method to improve hospital documentation and revenue.

Level of Evidence: Level IV

Study Type: Prospective cohort study with historical controls

**Key Messages:** What is already known on this topic: Documentation is critically important for hospital billing. Previous studies in trauma and acute care surgery have shown that large, intensive, and dedicated interventions can improve documentation and improve revenue generation.

What this study adds: By creating a list of Major Comorbidity/Complication (MCC) and Comorbidity/Complication (CC) codes for targeted review, two clinically busy trauma surgeons were able to perform limited, targeted chart review with significant improvement in DRG capture and Case Mix Index.

How this study might affect research, practice or policy: Improvements in documentation are facilitated by the targeted list of MCC and CC codes that other centers can use to improve their DRG capture, further demonstrating the financial benefit of a trauma and acute care surgery service.

## Introduction

In the early 1980s, the US Congress implemented a system of classifying inpatient hospital cases into diagnosis-related groups (DRGs) (1). DRGs are broadly broken into two groups: Medical and Surgical. The Centers for Medicare & Medicaid Services (CMS) assigns a relative weight for each Medicare Severity DRG (MS-DRG) (1–3). Case mix index (CMI) helps to even the playing field by accounting for clinical complexity, resource needs and hospital patient diversity that influence hospital reimbursement for Medicaid and Medicare patients (1–3). CMI is calculated by averaging the DRG weight for every patient discharged in a given time period. While Medicare assigns a base pay factor for each DRG, the payment each hospital receives per DRG weight on average will vary based on insurance contracts.

Each hospital thus has a unique Case Mix Associated Discharge (CMAD) value. The CMAD value (in dollars) is the amount of revenue a hospital receives per discharge and per DRG weight inclusive of all private and government insurance contracts.

Unfortunately, this system is only as accurate as the documentation that the clinicians chart. Other studies in the literature have demonstrated that intensive chart review in trauma surgical patients can increase DRG and revenue capture (4,5). These previous studies involved concurrent review either by a panel of surgeons or clinical documentation specialists increasing CMI. Unfortunately, these prior studies were limited to a trauma division in a single hospital. Further, such studies involved intensive chart review. We hypothesized that by looking for a small subset of diagnoses, a small number of surgeons could review cases across a multi-hospital integrated health system and generate a significant improvement in CMI, both for the trauma division and for all surgical patients across the system.

## Methods

### Intervention

Approval was obtained from the IRB at Baystate Medical Center. Starting in January 2024, two trauma surgeons reviewed the charts of patients cared for by trauma surgery attendings (TACS patients) twice a week. In October 2024, this was extended to select surgical patients at all 4 hospitals in the system who were noted to have length of stay (LOS) longer than the geometric mean LOS (GMLOS) or who were noted to have low value DRGs despite being in Intensive Care Unit (ICU) or stepdown. The working DRG, LOS, and GMLOS for all patients was displayed in a dashboard on a daily basis. Recommendations for improved documentation were emailed to the attending and resident or APP who wrote the note; the treating team made the final decision on what to document. These reviews focused on improving documentation for a subset of Major Comorbidity/Complication (MCC) or Comorbidity/Complication (CC) diagnoses (Table 1) that were identified based on limited chart review of surgical patients. In the full intervention period, chart reviews were performed 5 days a week by one of the two trauma surgeons, who were trained in coordination with the clinical documentation integrity (CDI) team to determine which diagnoses to focus on. This training consisted of a four-hour session to understand the mechanics of DRG capture, CDI review, and documentation support. Reviews took approximately 30 minutes per day. Chart review by the CDI support team was ongoing.

**Table 1:** Select Major Comorbidity and Complication (MCC) (bold) and Comorbidity or Complication (CC) diagnoses specifically included in this study to be documented when present.

| R6510-SIRS of non-infectious origin without acute organ dysfunction | **K66.1-Hemoperitoneum** |
|---|---|
| **R65.11-SIRS of non-infectious origin with acute organ dysfunction** | **R57.1-Hypovolemic Shock** |
| **A41.9-Sepsis** | D62-Acute posthemorrhagic anemia |
| **R65.20: Severe sepsis** | **K80.67 Calculus of gallbladder and bile duct with acute and chronic cholecystitis with obstruction** |
| **R65.21-Severe sepsis with septic shock** | **K65.0-Peritonitis** |
| R7881-Bacteremia | **K55.021-Acute infarction of small intestine** |
| E87.2-acidosis | **K85.10 Acute Biliary pancreatitis** |
| **S06.6X0A: Traumatic subarachnoid hemorrhage without loss of consciousness,** | **K83.09 Acute Cholangitis** |
| **S06.6X1A-Traumatic subarachnoid hemorrhage with loss of consciousness of 30 minutes or less** | **K55.019 Acute reversible ischemia of small intestine** |
| **S06.5X0A: Traumatic subdural hemorrhage without loss of consciousness** | **G92.9-Toxic Encephalopathy** |
| **S06.5X9A-Traumatic subdural hemorrhage with loss of consciousness of unspecified duration** | E87.1-hyponatremia |
| D68.32-Hemorrhagic disorder due to extrinsic circulating anticoagulants | **N17.0-Acute kidney failure with tubular necrosis** |
| **R40.20 - Traumatic Coma** | N17.9-Acute kidney failure |
| **G93.5- Brain Compression** | K76.6-Portal Hypertension |
| **I77.70-Dissection of unspecified artery** | E11.10-Type 2 diabetes mellitus with ketoacidosis without coma |
| S27.0XXA-Traumatic pneumothorax | **E43-Unspecified severe protein-calorie malnutrition** |
| **S27.2XXA-Traumatic hemopneumothorax** | Z68.41-Body mass index (BMI) 40.0-44.9, adult |
| S22.49XA-Multiple fractures of ribs, unspecified side, initial encounter for closed fracture | Z68.42-Body mass index (BMI) 45.0-49.9, adult |
| **J96.01-Acute respiratory failure with hypoxia** | **U07.1-COVID 19** |
| S22.20XA-Sternal Fracture | I48.20-Chronic atrial fibrillation |
| **S36.116A-Major laceration of liver** | **R57.0 Cardiogenic Shock** |
| **S36.032A-Major laceration of spleen** | **I21.A1 Type 2 MI** |
| T79.A29A-Traumatic Compartment Syndrome of Lower Extremity | **J81.0 Acute Pulmonary Edema** |
| **S14.0XXA-Concussion and edema of cervical spinal cord, initial encounter** | **K72.00 Acute Liver Failure** |
| **T79.4XXA-Traumatic shock, initial encounter** | E87.8 Refeeding Syndrome |

### CMAD Value

The value of a Case Mix Associated Discharge for the hospitals in the system was pulled from publicly available data from the Massachusetts Center for Health Information and Analysis from FY19-FY23 (6).

### Statistical Analysis

Statistical analysis was performed using R 4.4.0. The control group was all patients discharged with surgical DRGs across the 4 hospitals in the system from 10/2022 to 12/2023. The limited intervention group (TACS patients only) was 01/2024 to 09/2024. The full intervention group (surgical patients across the system) was 10/2024 to 07/2026. The primary outcome was surgical CMI. Secondary outcomes were CMAD value per case, overall LOS, and geometric mean LOS. Continuous outcomes were compared across the pre-, limited- and full-intervention time frames using ANOVA, with post-hoc Tukey-Kramer analysis to compare between pairs of periods. Comparisons between two groups, i.e. pre and limited/full for trauma patients, and pre/limited and full for non-trauma patients were performed using T-Test. A p<0.05 was considered significant.

## Results

The average CMAD value for the health system from FY19-FY23 was $13,467.29 (95%CI $13,145.88-$13,788.71). There were 13,807patients (916 TACS) in the pre- intervention group, 9,299 (582 TACS) in the limited intervention group and 22,453 (1538 TACS) in the full intervention group. In the limited and full intervention groups between 10-35 patients a day were reviewed, representing roughly 10% of surgical inpatients each day.

Surgical CMI increased significantly across the three periods (2.83 (95%CI 2.79- 2.87) to 2.86 (95%CI 2.81-2.91) to 2.96 (95%CI 2.93-3.00), ANOVA p <0.001; Figure 1). A Tukey-Kramer analysis demonstrated significant differences between full and limited periods (p< 0.001), full and pre periods (p<0.001), but not the pre and limited periods (p=0.880). CMAD value per case increased significantly ($38,140 (95%CI $37,612-$38,667) to $38,532 (95%CI $37883-$39,182) to $39,905 (95%CI $39,466-$40,344), ANOVA p <0.001). Tukey-Kramer analysis demonstrated significant differences between full and limited periods (p = 0.002), full and pre periods (p<0.001), but not pre and limited periods (p=0.643).

**Figure 1:**
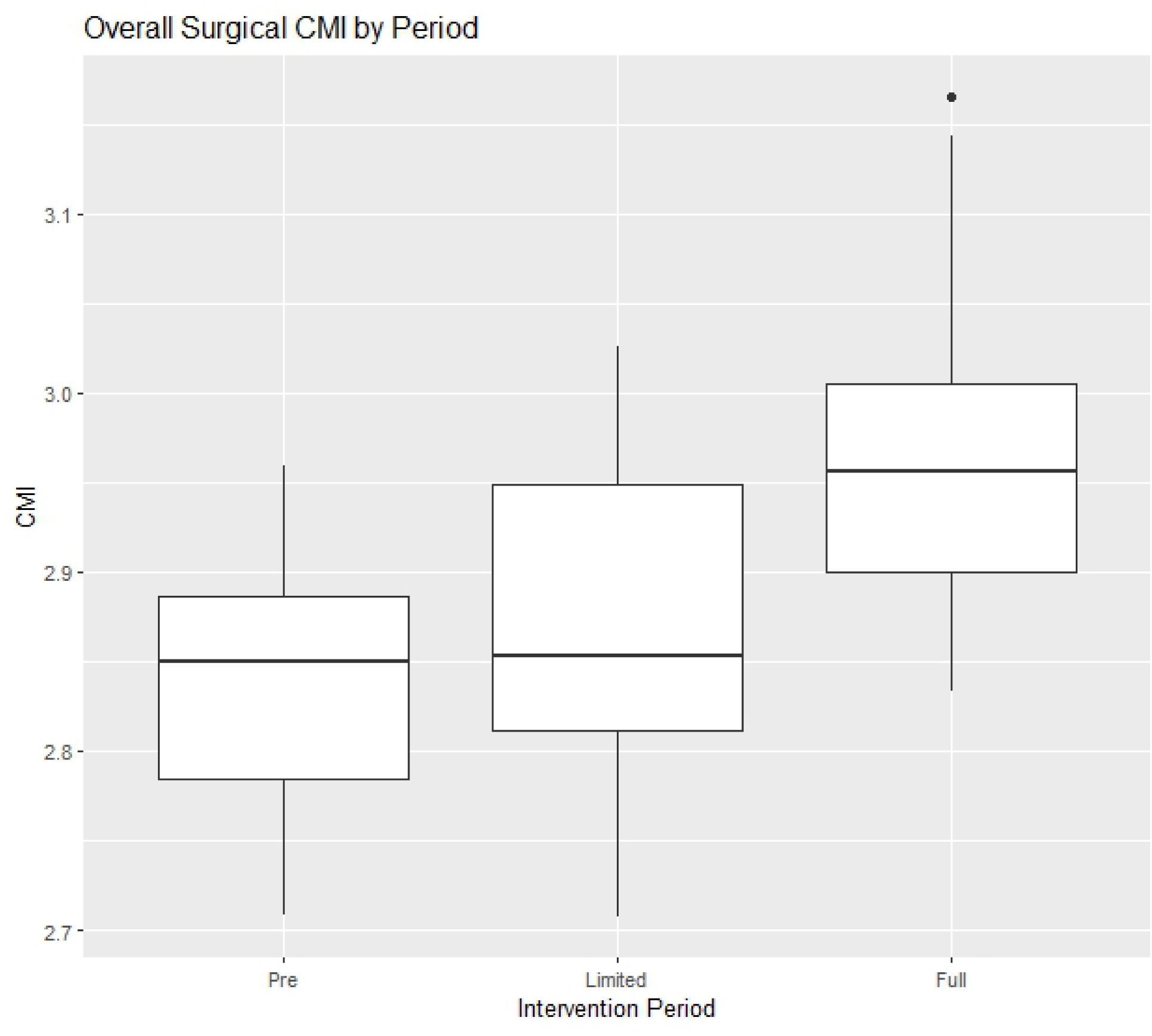
Increase in Surgical CMI across the four-hospital system with targeted chart review. Limited intervention focusing only on trauma patients, Full Intervention select surgical patients across the system.

For TACS patients alone, the significant increase in CMI occurred between the pre and limited/full phases (3.66 (95%CI 3.45-3.87) v. 3.95 (95%CI 3.81-4.09), p = 0.023) with no significant change between the limited and full phases (p = 0.987), For non TACS patients, the significant increase in CMI occurred between the pre/limited and full phase (2.78 (95%CI 2.75-2.81) v. 2.89 (95%CI 2.86-2.92), p<0.001) with no significant change between the pre and limited phases (p = 0.903) (Figure 2).

**Figure 2:**
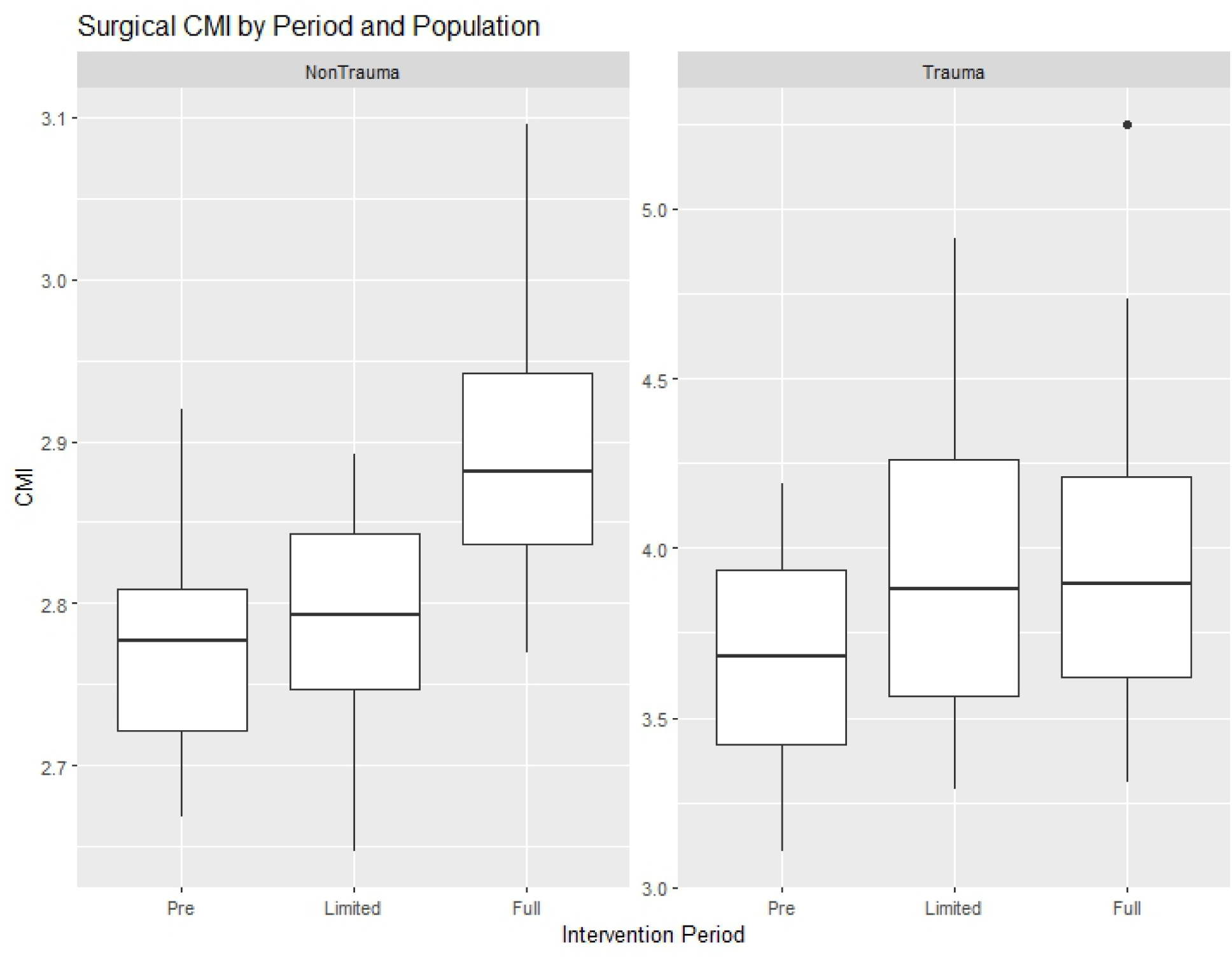
Surgical CMI increase by intervention period divided up by Trauma patients and NonTrauma Patients.

Overall LOS decreased significantly across the three time frames (6.52 days (95%CI 6.35-6.69) to 6.18 days (95%CI 5.99-6.36) to 6.29 days (95%CI 6.17-6.41), p = 0.016).

Tukey-Kramer analysis demonstrated significant difference between the Limited and Pre time frame (p = 0.020), but not Full and Pre (0.060) or Full and Limited (0.629). Geometric mean LOS (expected LOS for a patient’s DRG) changed significantly (4.46 days (95%CI 4.41-4.52) to 4.39 days (95%CI 4.32-4.45) to 4.50 days (95%CI 4.46-4.54), p=0.019).

Tukey-Kramer analysis demonstrated significant differences between Full and Limited periods (p = 0.014), but not between Limited and Pre periods (p = 0.161) or Full and Pre periods (0.612).

## Discussion

The institution of targeted chart review by trauma surgeons significantly increased surgical CMI across the system by 4.6% from baseline. Importantly, these reviews were performed in addition to and in collaboration with the CDI team chart reviews which were ongoing across all three periods. Direct facility revenue data were not available for all patients, but hospital inpatient revenue is directly tied to total DRG weight via the Case Mix Associated Discharge (6). Thus, we estimated the revenue increase as the difference in mean CMAD value per case between the full intervention and pre-intervention periods ($39,905 vs. $38,140), or $1,765 per patient for all surgical patients across the system once fully rolled out.

Although it is possible that the CMI increase was due to more severely ill patients being admitted in the study period, there was no change in observed overall length of stay. Because geometric mean LOS is derived from the assigned DRG, its increase reflects a change in documented severity rather than a change in how long patients actually stayed. The CMI increase thus likely reflects, at least in part, improved documentation capture rather than a change in patient complexity, although LOS is an imperfect surrogate for acuity. These findings are consistent with other studies in the literature investigating improvement in documentation in improve CMI. Some studies in trauma patients have reported somewhat larger CMI increases of 13.7% and 17.2% utilizing more reviewers(4,5). A similar study focusing on neurotrauma patients and templated documentation demonstrated 13-22% increase in reimbursement for these patients (7).

Whereas a study focusing on vascular surgery patients demonstrated an increase in CMI of 5.6% (8). We were able to accomplish a similar CMI increase across all surgical patients in 4 hospitals in the integrated system. In addition, the scale of our intervention was an order of magnitude larger. While the other studies in the literature range between 400-1800 patients per year, our intervention affected over 12,000 surgical patients per year (5,7).

The improved clinical capture was achieved by focusing on a set of high value diagnoses that are frequently missed. Because the reviews specifically targeted CC and MCC diagnoses, an increase in CMI was the expected direction of effect; the question we specifically addressed is whether a limited, targeted effort by a small number of surgeons produces a measurable change at the scale of a multi-hospital system.

Given the complexity of clinical care amongst surgical patients, a focus on improving documentation to accurately portray clinical severity is essential. CDI programs have a role in this sector and a few publications denote the impact of an organized CDI team for trauma systems (5). The surgeon review we performed here is intended to complement the CDI team rather than replace it. Here, two trained surgeons worked in collaboration with the CDI team, bringing added value because of the clinical knowledge and understanding they had over and above the nonclinical members of the CDI team. While CDI teams may improve over time, the leadership and experience of a clinically active surgeon is indispensable to ensuring diagnoses are captured that may be documented in a nonstandard way. Our study shows that such reviews are not only feasible by a small number of trained physicians but can have a large positive effect on surgical CMI and potentially revenue across a system, with a time commitment of approximately 30 minutes per day. Because this benefit accrues primarily to the hospital through facility reimbursement rather than to individual surgeons, surgeon engagement in such reviews may depend on the compensation model and on institutional support for reviewer time. In addition, while a trauma surgeon’s time is valuable, the potential increase of $1765 per patient in a system with over a thousand surgical patients per month provides adequate justification for compensation commensurate with the surgeon’s time. We therefore suggest that such reviews be compensated with administrative FTE. Further, this additional revenue to the hospital benefits the Department of Surgery by improving operating margin and allowing the availability of more resources to individual surgeons.

There are some limitations to our study. Unfortunately, due to the timing of data availability, revenue cycle data were not available for all patients. However, we can calculate the monetary value of the improved CMI using the publicly available value of CMAD for the hospitals in our system which is an average value of revenue per discharge per unit of DRG weight. Additionally, our study is limited by the fact that it was all done in one healthcare system. This was a pre/post design without a concurrent control group, so we cannot exclude other changes during the study period, such as shifts in service lines, case volume, or coding practices, as contributors to the CMI increase. Patient acuity was also not measured independently of DRG-based measures. Finally, the CMAD value was derived from FY19-FY23 data and may not reflect reimbursement during the full study period (10/2022-7/2026). Because CMAD is an average across all payer contracts, the revenue estimate does not account for payer mix, denials, or uncollected payments, and actual collections may be lower.

In conclusion, having surgeons review targeted surgical patients for high value documentation capture significantly enhanced the case mix index and increased estimated revenue by $1,765 per surgical patient across a multi-hospital integrated health system. This improvement in CMI and estimated revenue likely reflects enhanced documentation rather than increased patient illness severity. Such documentation reviews may represent a practical method to improve hospital documentation and revenue.

These data were presented at the annual meeting of the Rocky Mountain Trauma Society in Aspen, CO on 1/20/2025.

## Data Availability

All data produced in the present study are available upon reasonable request to the authors

